# Systematic comparison of Mendelian randomization studies and randomized controlled trials using electronic databases

**DOI:** 10.1101/2022.04.11.22273633

**Authors:** Maria K. Sobczyk, Jie Zheng, George Davey Smith, Tom R. Gaunt

## Abstract

Mendelian Randomization (MR) uses genetic instrumental variables to make causal inferences. Whilst sometimes referred to as “nature’s randomized trial”, it has distinct assumptions that make comparisons between the results of MR studies with those of actual randomized controlled trials (RCTs) invaluable. To scope the potential for (semi-)-automated triangulation of MR and RCT evidence, we mined ClinicalTrials.Gov, PubMed and EpigraphDB databases and carried out a series of 26 manual literature comparisons among 54 MR and 77 RCT publications. We found that only 11% of completed RCTs identified in ClinicalTrials.Gov submitted their results to the database. Similarly low coverage was revealed for Semantic Medline (SemMedDB) semantic triples derived from MR and RCT publications –25% and 12%, respectively. Among intervention types that can be mimicked by MR, only trials of pharmaceutical interventions could be automatically matched to MR results due to insufficient annotation with MeSH ontology. A manual survey of the literature highlighted the potential for triangulation across a number of exposure/outcome pairs if these challenges can be addressed. We conclude that careful triangulation of MR with RCT evidence should involve consideration of similarity of phenotypes across study designs, intervention intensity and duration, study population demography and health status, comparator group, intervention goal and quality of evidence.

## Introduction

Randomized controlled trials (RCTs) are deemed the “gold standard” in evaluating the efficacy of interventions and guiding practice in clinical research, with well-established methodology^1^. In RCTs, a selection of individuals intended to represent the target population is randomly assigned to a treatment or control group, allowing estimation of the intervention’s effectiveness in the absence of confounding variables and reverse causality that are present in observational studies. In the past two decades an approach to causal inference using natural genetic variation, known as Mendelian Randomization (MR) – usually implemented as an instrumental variable (IV) analyses – has gained popularity^2,3^. This approach has been referred to as “nature’s randomized trials”^4^, and is based on the randomization from parents to offspring of genetic variants encapsulated in Mendel’s laws of segregation and independent assortment^2,5^. At a population level the randomization is approximate, but still allows genetic variants that are robustly associated with the measured exposure to be used to estimate the unbiased causal effect of an exposure (generally acting across life) on health outcomes, as long as certain assumptions, discussed in detail elsewhere^2,3,6^, are met.

Despite drawing on observational data, the MR approach broadly aligns with that of a randomized controlled trial, where the goal is to estimate the causal effect of an intervention on the given endpoint based on groups (arms) which do not differ with respect to confounding variables (Figure 1). However, since in MR randomization takes place at conception, the time lag to the start of outcome recoding is longer^7^ compared to RCTs, where median duration of phase 3 trials is 40 months^8^. Similarly to RCTs, most MR analyses should be free of confounding and reverse causation bias due to variants being allocated randomly before birth and outcome condition onset.

**Figure 1.**
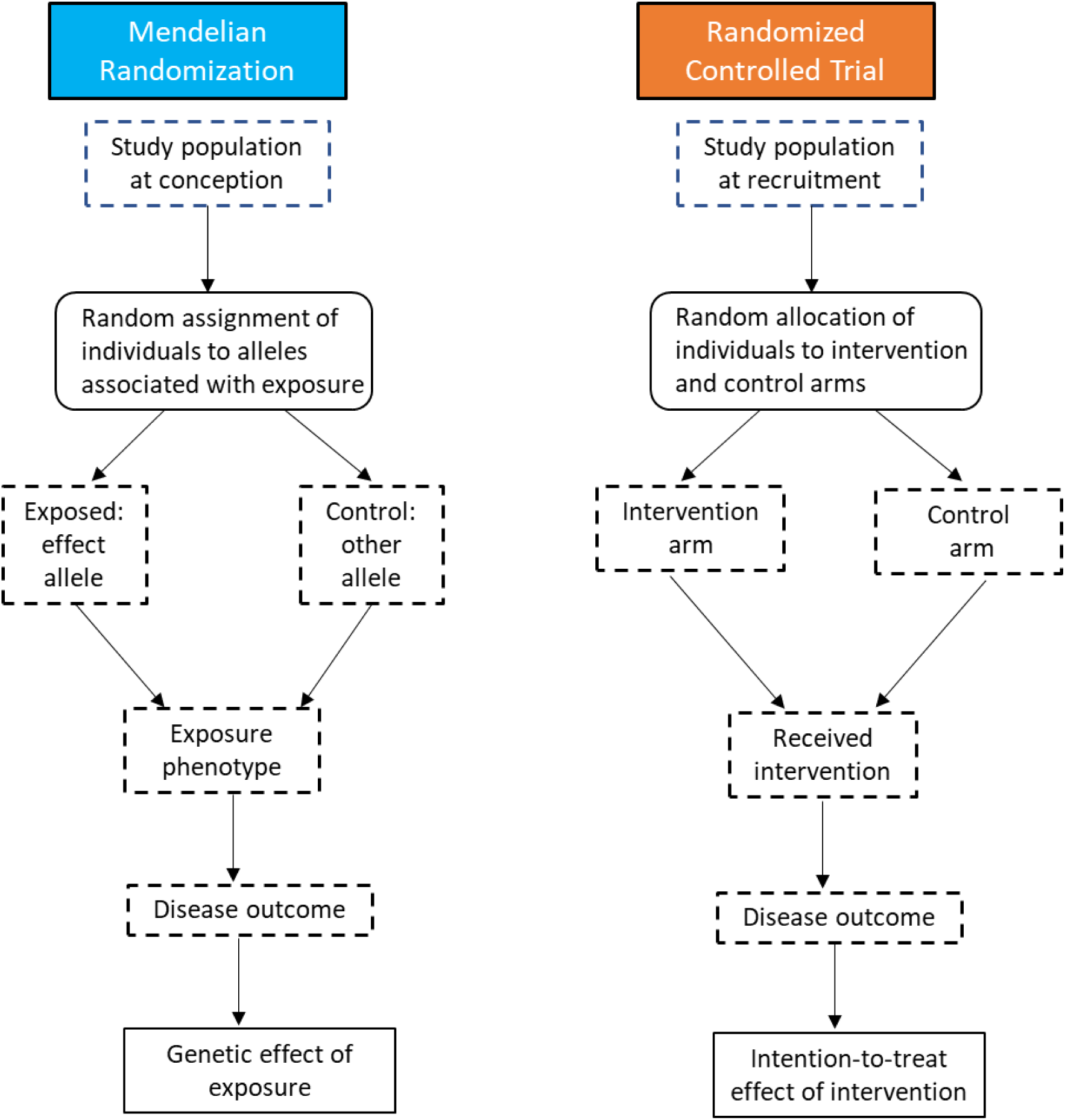
Comparison of Mendelian Randomization and Randomized Controlled Trial design. After: Nitsch *et al*. (2006), Ebrahim & Smith (2008), Ference (2018).

Previous research has shown examples of evidence triangulation where MR results predicted the overall RCT results based on totally orthogonal data with an unrelated set of systematic errors and biases^9^. For instance, MR demonstrated the lack of effect of genetically-predicted concentrations of HDL-C on cardiovascular events^10–12^ as well as selenium in prostate cancer prevention trials^13,14^. On the other hand, MR showed the beneficial effect of lifelong endogenous low LDL-C levels^15^, HMG-CoA reductase inhibition (statin drug target) and PCSK9 inhibition on cardiovascular disease^15,16^, while predicting also the increased risk of type 2 diabetes (T2D) as a side-effect of statin usage. However, three independent MR studies were at odds with later RCTs by predicting increased risk of T2D also as a side-effect of PCK9 inhibition^17^.

There are several possible explanations for apparent or real discordance in the results of RCT and MR studies. These range from different durations, magnitude and time-varying nature of the exposure, origin of the study populations, and natural genetic variation imperfectly mimicking the molecular action of the drug, some of which we explore. The direct comparison of MR and RCT findings is facilitated by the use of a precisely-defined estimand^18^, for example, the effect on incident coronary heart disease risk of lowering LDL cholesterol by 1 mmol/l for 5 years. Whilst RCTs will estimate something close to this, and be scalable to it, with MR studies the exposure difference associated with the genetic instruments will often exist from birth (or before) and may change in magnitude over time^19^. This is discussed further in the Supplementary Box.

Whilst RCTs can provide the highest-quality evidence, they may have limitations. They are often expensive to carry out, can be of small size and lack external validity^20^, have short follow-up and typically take place after disease onset^21,22^. As in other study types, RCT results may be flawed due to poor design and execution, e.g. imperfect randomization, unblinding and differential loss to follow-up between study arms.

Unlike RCTs, MR studies are inexpensive and quick to perform when suitable genetic instruments are available. Therefore, they can potentially prioritize intervention-condition pairs to assess in RCTs. Moreover, it has been proposed that MR also guide the design of RCTs, improving eligibility criteria to prioritise groups most likely to benefit, suggesting diseases for composite endpoint construction, and alerting to potential side-effects^1,23^. Since MR analyses suffer a different set of biases than RCTs, MR evidence can be used to complement RCTs and other study designs in the triangulation framework to guide therapeutic development and clinical practice^24–26^. Finally, the extensive use of existing observational data for MR enables intervention targets to be evaluated in a wider range of sub-populations than is feasible for RCTs (improving generalizability), and allows comparisons to be made that might be unethical in experimental studies, for example when there is strong evidence in favour of a particular treatment.

The goal of this research is to survey the extent of concordance between MR and RCT studies to date and identify possible factors for disparities in the direction of effect, which limit the ability to extrapolate from MR results to RCTs and increase the complexity of the triangulation process. In this study, we aimed to carry out a systematic analysis of MR and RCT results using automated mining of data in the public domain, including the ClinicalTrials.Gov^27^, EpigraphDB^28^ and PubMed databases. We evaluate the comprehensiveness and scope of the data available and potential for comparative analyses between MR and RCTs. We then go on to develop a series of case-studies looking in detail at MR and RCT comparisons across 26 exposure-outcome pairs. Throughout, we use the term “intervention” as synonymous with “exposure” and “condition” as synonymous with “outcome”.

## Results

### ClinicalTrials.Gov data overview

In total, we found 379,094 individual studies were registered with a unique ClinicalTrials.gov identifier. We filtered them using a number of steps to identify RCTs and facilitate comparison with MR (Figure 2). In our analysis, we identified 166,954 RCT studies (44% of the total). To allow semi-automated comparison with MR studies, we focussed on the study subset which submitted their statistical analysis results to the database (referred to as the **main dataset**, Supplementary Dataset 1). However, we found that only 4% of studies – 13,807 met this criterion, along with including background information on the trial. To expand that number, we also considered an additional 23,080 RCT studies which did not publish their results in ClinicalTrials.gov but instead linked to a peer-reviewed publication (referred to as the **literature dataset**, Supplementary Dataset 2).

**Figure 2.**
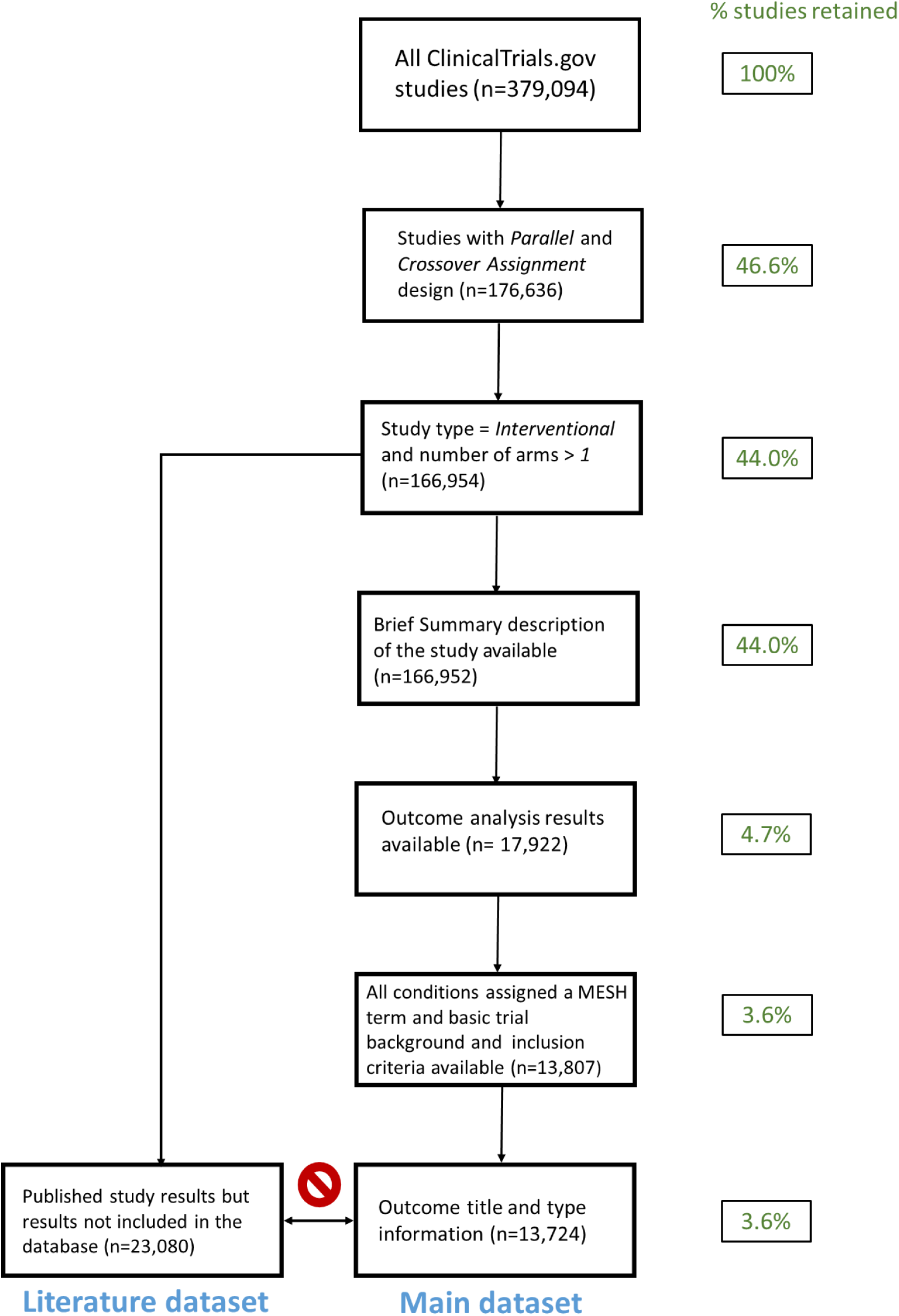
Filtering steps applied to ClinicalTrials.Gov database. Filtering was designed to identify RCT trials whose final results statistics were uploaded to the database (*Main dataset*). In addition, other RCTs which published their findings in scientific journals were identified (*Literature dataset*).

The majority of RCTs in the main dataset followed parallel assignment of participants to treatment (Figure S1a), most were designed for treatment (n=10,812, Figure S1b), rather than prevention (n=1,422) and the vast majority of them had been completed (Figure S1c). More trials were observed to be in phase 3 than 4 (Figure S1d), most trials included both males and females (Figure S1e), and a great majority had 2 arms (Figure S1f). The median number of primary outcomes was 1 (Figure S2a), with a median of 5 secondary outcomes (Figure S2b). Over half of studies report at least 1 result with *p*-value less than 0.05 (Figure S2c). Comparison with features of all RCTs in the database showed that our selection was broadly representative (Supplementary Dataset 3), although our dataset was enriched for completed and late-phase trials.

### Suitability of MeSH annotation

In order to attempt automated matching of RCTs and MRs involving similar interventions and outcomes for RCTs and MR, we needed to first establish the quality of annotation of RCTs with MeSH (Medical Subject Headings) in ClinicalTrials.Gov. The most common intervention was Drug (Table S1). Since we were only interested in the intervention types which can be instrumented by MR, we also focussed on the 4^th^ and 7^th^ most popular types of interventions: Behavioral and Dietary Supplement. We found that MeSH intervention annotations were missing for only 19% and 16% of Drug interventions in the Main and Literature datasets, accordingly (Table 1). However, the overwhelming majority of RCTs in the Behavioral and Dietary Supplement category did not contain a MeSH intervention term. Due to well standardised disease taxonomy, a much lower level of missing data was found for MeSH condition terms. This allowed us to proceed with automated analysis of Drug RCT data, however, for Behavioral and Dietary Supplement we were only able to do a manual screening for RCTs with corresponding MR studies.

**Table 1.** Completeness of MeSH term annotation amongst the chosen intervention types in the *Main* (RCT results available in ClinicalTrials.Gov) and *Literature* (RCT results unavailable in ClinicalTrials.Gov but study linked to a publication with results) datasets.

| Dataset | Intervention type | Total | intervention<br>MeSH<br>missing | %<br>missing | condition<br>MeSH<br>missing | %<br>missing |
| --- | --- | --- | --- | --- | --- | --- |
| Main | Drug | 10,187 | 1,915 | 18.8% | 936 | 9.2% |
| Literature | Drug | 9,554 | 1,526 | 16.0% | 914 | 9.6% |
| Main | Behavioral | 1,013 | 872 | 86.1% | 203 | 20.0% |
| Literature | Behavioral | 2,427 | 2,248 | 92.6% | 496 | 20.4% |
| Main | Dietary Supplement | 215 | 102 | 47.4% | 30 | 14.0% |
| Literature | Dietary Supplement | 662 | 449 | 67.8% | 118 | 17.8% |

### Pharmaceutical interventions in RCTs and MR

Genetic instrumental variables in MR can be used as proxies for pharmaceutical interventions in RCTs. Protein (pQTL) or expression (eQTL) quantitative trait loci (QTL), i.e. variants associated with expression of protein drug targets are used to directly proxy the action of a drug. Here we use the biggest MR dataset for drug target protein-disease associations, examined in whole blood, from Zheng et al. (2020)^29^. We focussed on *cis*-acting instruments as a more specific marker for drug efficacy as *trans-* instruments are more likely to be pleiotropic, potentially leading to spurious results^23^.

We matched the drug target proteins in Zheng et al. (2020) with drug-gene associations sourced from EpigraphDB^28^. This allowed us to merge the Zheng et al. (2020) dataset with the main and literature RCT dataset via the drug listed in EpigraphDB and MeSH drug intervention term, accordingly. For the outcome, we were then able to match RCTs and MR manually due to the reasonably low number of hits. The results, displayed in Table 2 show overlap of the RCT and MR datasets. We found 4 drugs: evolocumab/alirocumab, ustekinumab and mipomersen that share support from both MR and RCT studies. evolocumab/alirocumab and mipomersen inhibit key players (PCSK9 and apoB) in lipid transport helping to lower plasma LDL-C levels^30^. The Zheng et al. (2020) MR study showed a negative effect of reduced PCSK9 levels on high cholesterol in the UK Biobank, while in 25 RCT studies drug-induced abrogation of PCSK9 activity led to positive outcomes in the treatment of hyperlipidemia, hypercholesterolemia and dyslipidemias in general. Similarly, reduced expression/activity of apoB in MR and 6 RCT studies resulted in genetically predicted lower levels of LDL cholesterol and total cholesterol in the UK Biobank as well as improved outcomes in the treatment of dyslipidemias, respectively. The third example of a good match between RCT and MR studies concerns inhibition of the p40 subunit of interleukin 12 and 23 (IL12B)^31^. Both MR and 21 RCTs show benefit of inhibition of p40 on immune-mediated disease: psoriasis and inflammatory bowel disease.

**Table 2.** Drug target-disease matches supported by evidence from MR (blood pQTL instruments, Zheng et al. 2020) and RCT studies (*Main dataset* from ClinicalTrials.Gov).

| MR Exposure | RCT Drug Intervention | MR Outcomes | RCT Conditions | Matching trials | Concordant direction of effect? | xQTL |
| --- | --- | --- | --- | --- | --- | --- |
| PCSK9 | EVOLOCUMAB, ALIROCUMAB | Non-cancer illness code self-reported: high cholesterol \|\| id:UKB-a:108 | Hyperlipidemia, Dyslipidemia, Hypercholesterolemia, Mixed Dyslipidemia | 25 | Yes | pQTL |
| APOB | MIPOMERSEN | LDL cholesterol \|\| id:300, HDL cholesterol \|\| id:299, Triglycerides \|\| id:302, Non-cancer illness code self-reported: high cholesterol \|\| id:UKB-a:108, Total cholesterol \|\| id:301, | Hyperlipidemia, Dyslipidemia, Hypercholesterolemia, Mixed Dyslipidemia | 6 | Yes | pQTL |
| IL12B | USTEKINUMAB | Non-cancer illness code self-reported: psoriasis \|\| id:UKB-a:100; Ulcerative colitis \|\| id:970; Crohn's disease \|\| id:12; Inflammatory bowel disease \|\| id:294 | Psoriasis, Psoriatic Arthritis, Crohn's Disease, Colitis, Inflammatory Bowel Disease | 21 | Yes | pQTL |

In general, pQTL MR-based prediction of drug target-condition pairs offered good recall when compared with the pairs in the Open Targets Platform for the proteins with MR evidence. The only drug target indications missing included conditions not analysed in the MR study, such as CD33 protein being the drug target for treatment of leukemia, with the exception of ACHE (acetylocholinoesterase) whose inhibitors (galantamine, donepezil, rivastigmine) are used for treatment of cognitive decline in Alzheimer’s disease^32^.

We also compared the RCT dataset with Zheng et al. blood transcript expression (eQTL)-derived MR analysis (available in EpigraphDB: https://epigraphdb.org/xqtl). In total, we identified 15 drug target-disease matches in the eQTL dataset (Table S2), although unlike in the pQTL matches, the direction of effect in MR was incorrect in 8 cases. Nevertheless, the eQTL MR results agreed with some well-known drug effects: HDL-C and LDL-C lowering action of CETP and HMGCR inhibitors^33^, respectively, and blood pressure-lowering action of ACE inhibitors^34^.

### PubMed-sourced MR and RCT studies

In addition to searching through the ClincalTrials.Gov database, we also queried PubMed for RCT and MR publications. In total, we found 3,538 MR studies published since mid-2000s and 63,187 RCTs published since 1970 until 2020 (Figure 3).

**Figure 3.**
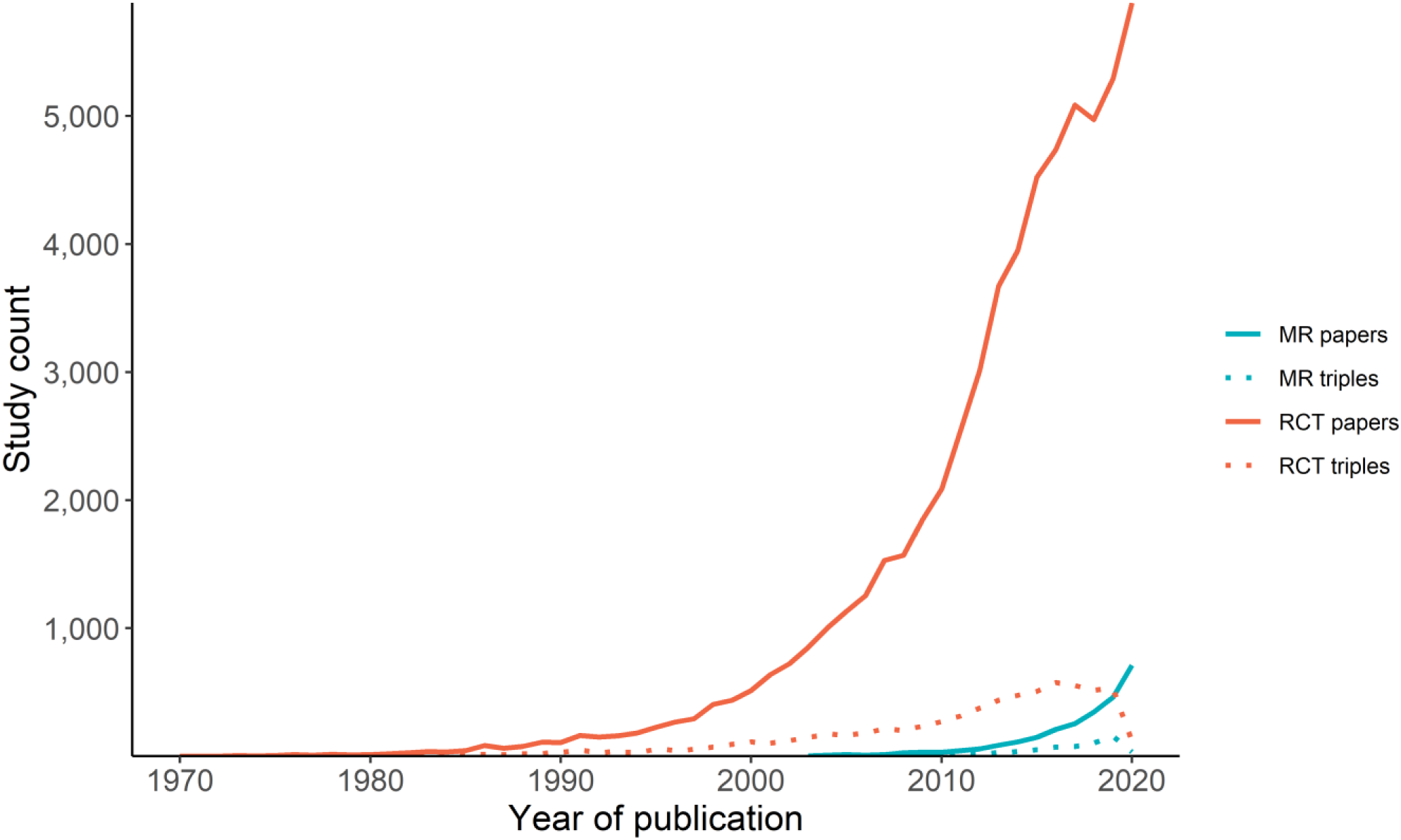
Popularity of MR and RCT studies over time. We compare counts of MR and RCT papers indexed by PubMed (solid lines) with number of semantic triples derived from them using SemMedDb (dashed lines).

### Semantic analysis with SemMedDB

We subsequently wanted to establish the thematic overlap between MR and RCT studies using an alternative method involving semantic analysis. SemMedDB^35^ provides a vast repository of semantic predications (subject-predicate-object triple, e.g. LDL-C causes ischemic heart disease). We linked the MR and RCT publications identified by our PubMed search to their corresponding SemMed triples in EpigraphDB using PubMed ID. Overall, only 12% and 25% of RCT and MR papers, respectively, had a semantic triple associated with them (Figure 3). When ignoring the predicate, and focusing only on the subject and object, we found a total of 10,875 unique exposure-outcome pairs, discussed in detail in the Supplementary Note. However, only 125 of these were found to be shared across MR and RCT studies.

We then investigated the 125 matching subject-object pairs between MR and RCT studies (Table S3), as well as individual top counts among subjects (Table S4) and objects (Table S5). T2D, insulin and obesity were found among the top shared risk factors, along with lipids and vitamin D. Top outcomes included T2D, cardiovascular disease, asthma and Alzheimer’s disease.

### Case studies of matching MR and RCTs

Since our semi-automatic mining of MR and RCT literature brought limited results for behavioural and nutritional interventions, we selected 26 intervention-outcome case-studies by manual mining of the literature representing common lifestyle risk factors, dietary and behavioural exposures, paired with common cardiovascular, glycemic, neuropsychiatric, musculoskeletal, autoimmune and cancer outcome phenotypes. In total, we surveyed 54 MR and 77 randomized controlled trial publications (RCTs and meta-analysis of RCTs, Supplementary Dataset 4, Figure 4) which were systematically compared across several criteria shown in sample Table S6, and encompass those in the popular PICO (Population, Intervention, Comparison, and Outcome) framework^36^.

**Figure 4.**
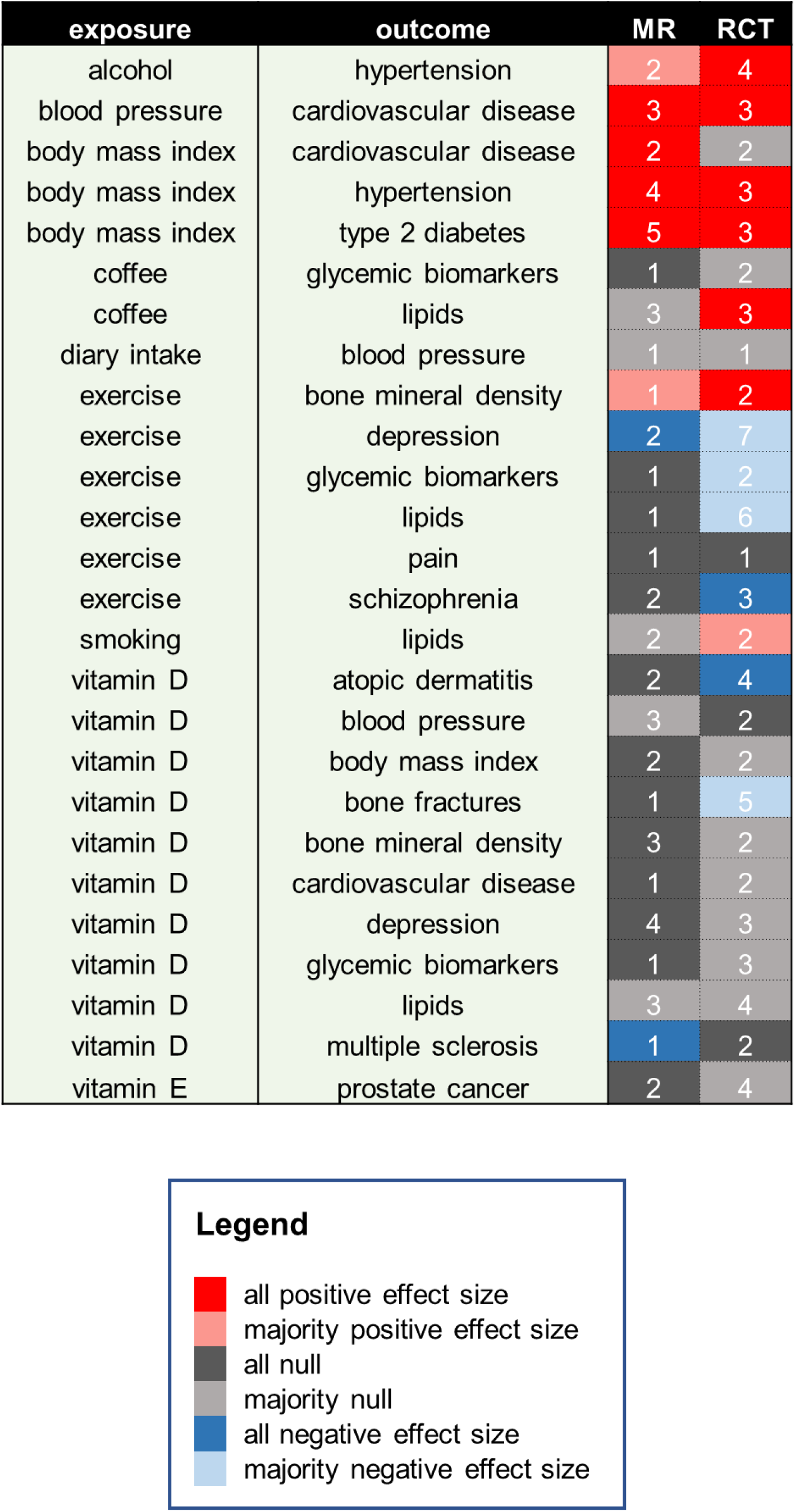
Summary of case series of MR and RCT studies with matching exposures (interventions) and outcomes (conditions). The values correspond to the number of analysed studies in a given category, while the cell background colour indicates summary direction of effect on the outcome when exposure is increased – we report direction of effect found either in all analysed studies or their majority (>50%).

There, we compare an MR study and two RCT meta-analyses on the effect of vitamin D supplementation in multiple sclerosis (MS)^37–39^. Whilst the RCTs looked at potential therapeutic effect of vitamin D in diagnosed MS patients over 6 months-2 years: measured disability (EDSS score) and recorded relapses as outcomes, the MR study took place in the general population and measured the causal effect of genetically predicted lifetime circulating vitamin D concentrations on prevention of MS. The conclusions of MR and RCT studies did not align well, with MR analyses providing evidence for reduced risk of MS conferred by higher vitamin D levels, but no significant therapeutic effect of vitamin D in existing MS was found in the 5 small meta-analyzed trials. Differences which may impact on the ability for MR to complement RCT studies are summarised in Table 3 and discussed below based on this series of case-studies.

**Table 3.**
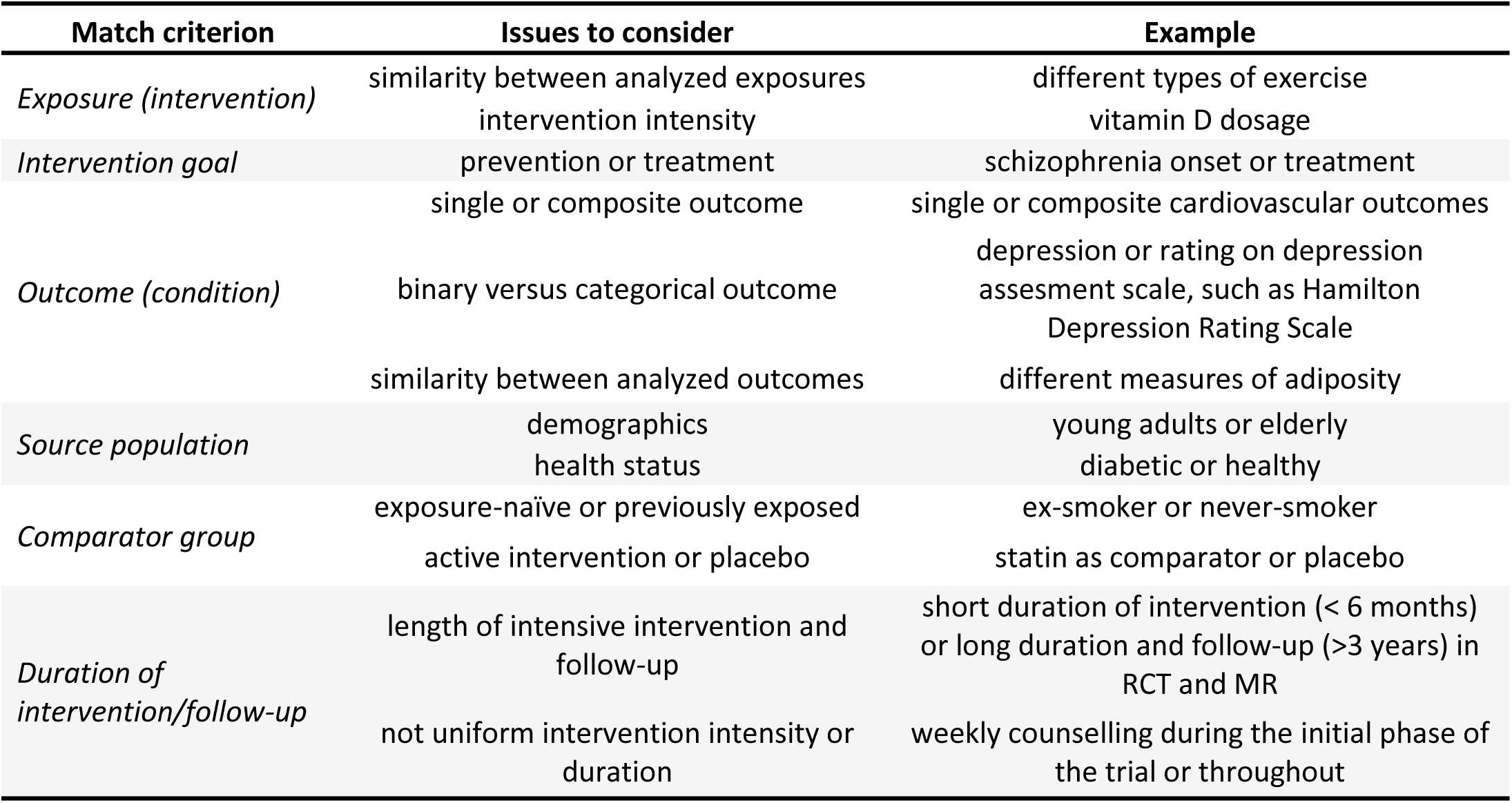
Overview of discussed criteria for assessment of alignment of MR and RCT study features.

### 1) Exposure/Intervention

We found that overlapping MR and RCT interventions are often not perfectly identical which may impact on the estimated direction of effect. For example, MR exercise exposures are based on genetic variants associated with self-reported physical activity (moderate-to-vigorous and vigorous)^40,41^ in studies assessing the effect on both lipids and bone mineral density (BMD). However, the corresponding RCTs used particular types of exercise, such as walking^42^, aerobic exercise^43,44^, progressive resistance training^45^ and maximal strength training^46^ as interventions. While a MR study^41^ and two trials^46,47^ showed concordant (Figure 4), positive effect of exercise on BMD, we found that the effect of exercise on lipids did not match between MR and RCTs, with MR study^40^ reporting null effect and trials generally finding positive effects on HDL-C concentration and negative on LDL-C, total cholesterol and triglycerides blood levels^43–45,48^.

Furthermore, intensity of intervention can affect the comparative value of MR and RCT study conclusions. The MR study of vitamin D levels on bone fractures^49^ was only able to assess linear effects of the normal range of circulating 25-hydroxyvitamin D concentrations. Consequently, the positive effect of high-concentration vitamin D (≥700 IU daily) on bone fractures in the elderly seen in RCTs^50–52^ may not have been accessible in the MR study.

### 2) Intervention goal

The intervention goal between MR and RCT studies can match (both prevention or treatment) or be misaligned which can potentially impact the ultimate conclusions of the study. We found the latter to be the case for the effect of exercise on schizophrenia. Two MR studies found a null preventative effect of exercise on schizophrenia^53,54^, while 3 meta-analyses of RCTs found a consistent effect of a variety of exercise types on improving total and negative symptoms of schizophrenia^55–57^ (Figure 4).

### 3) Outcome

The short duration of RCTs mean some outcomes (e.g. myocardial infarction) do not accumulate enough events to detect a significant effect, therefore composite measures grouping related diseases are often used. When comparing the effect of systolic blood pressure (SBP) on cardiovascular disease (CVD) outcomes, we found matching conclusions with elevated SBP increasing the risk of CVD both in RCT^58–60^ and MR^61–63^ studies (Figure 4), with MR studies using both single disease outcomes and a composite outcome. However, MR studies analysing the impact of BMI on cardiovascular disease found reduced adiposity led to reduction in arterial hypertension, CVD and stroke^64,65^, which contrasted with the results of one of the biggest RCTs to date. The Look AHEAD RCT in older type 2 diabetics found no preventative effect of weight loss on a composite outcome relating to mortality from cardiovascular causes, non-fatal myocardial infarction, non-fatal stroke or hospitalization for angina^66^.

Secondly, RCT outcomes, are often on a quantitative scale measuring symptom strength according to established metrics, (e.g. depressive symptoms on Hamilton Depression Rating Scale^67^). However, the best disease GWAS used to identify MR instruments often represent binary disease outcomes, which could potentially lead to differential conclusions due to reduced power to detect subtler therapeutic effects. While exercise is causally associated with reduced depression and depressive symptoms both in MR^68,69^ and RCTs^55,67,70–73^, the differences in outcome phenotypes could potentially contribute to null MR results^74,75^ and positive effect of vitamin D on attenuating eczema symptoms in RCTs^76–79^.

### 4) Source population

MR studies are likely to draw from a wider demographic than RCTs due to use of biobanks and GWAS consortia, while RCTs focus on high risk groups^23^. For example, while in the MR study conducted in general population, there was no strong significant effect of exercise on glycemic markers: HbA1c, fasting glucose and HOMA-IR^80^, a significant reduction was found in the meta-analysis of 32 RCTs involving patients with T2D^81^.

On the other hand, 5 Mendelian randomization^82–86^ studies along with 3 large RCTs^87–89^ consistently provide evidence that weight loss is causally associated with reduced risk of T2D (Figure 4), despite MR including the general population and RCTs focussing on at-risk individuals with impaired glucose tolerance.

As another example of possible demographics-driven differences in trial and MR results, MR studies on the relationship between vitamin D levels and atopic dermatitis were conducted in the general population^74,75^, while RCTs were conducted separately in children in Mongolia^76^ and Boston, USA^77^ with winter atopic dermatitis and in adolescent and adult Iranians^78,79^.

### 5) Comparator group

Firstly, due to ethical considerations, trials of harmful behaviours such as alcohol drinking and smoking focus on cessation or reduction in existing users, and do not include never-smokers or never-drinkers as controls, unlike MR studies, which can potentially lead to differences in effect^23^. Nevertheless, the two outcomes analysed here: hypertension for alcohol intake and lipids for smoking showed generally congruent results across study types (Supplementary Dataset 4).

Secondly, where it would be unethical to withhold already available efficacious treatments, trials will often include another active intervention in the comparator group^23^, for example statins in the trials of effect of PCSK9 inhibitors^90,91^ on LDL-C and cardiovascular events. Such RCT design can be mimicked by factorial MR estimating the interaction of multiple exposures, as shown in matching results of the equivalent MR study^16^.

### 6) Duration of intervention

While the magnitude of effect seen in trials with long (> 3 years: weight loss to treat hypertension^92,93^/type 2 diabetes^87–89^, blood pressure reduction to lower cardiovascular disease risk^58–60^) and short (< 6 months: alcohol intake reduction to lower blood pressure^94– 97^, exercise to benefit bone mineral density^46^/depression^55,67,71–73,98^) intervention may vary, we find both can result in directional effects consistent with MR results (Figure 4), although with exceptions^66,99^.

### Triangulation of MR and RCT results

Combining RCT and MR results can offer complimentary evidence on the effectiveness of interventions. Powerful examples include congruence of positive effect of high BMI on hypertension across MR^100–103^ and RCT^92,93^ studies, high BMI on T2D risk in MR^82–86,104^ and RCTs^87–89^, and the null effect of vitamin D on various glycemic markers in diverse populations in MR^105^, RCTs^106,107^ and RCT systematic review^108^.

We also found cases, where the majority of studies pointed to one direction of effect, with one MR or RCT identified as an outlier study. In these cases, having a wide array of MR and RCT studies (ideally meta-analysed) can be helpful in establishing the likely true causal direction of effect. For instance, 2 MR studies^109,110^, a meta-analysis of 5 RCT studies^111^ and two RCTs^112,113^ indicate no effect of vitamin E on prostate cancer incidence with one outlier RCT^114^ showing benefit of vitamin E supplementation in older smokers. Similar contrary findings were found for 1 RCT^115^ showing beneficial effect of vitamin D on preventing depressive symptoms, as opposed to null effect in 4 MR studies^116–119^ and 2 RCTs^120,121^.

On the other hand, MR analyses can show spurious disagreement with the rest of the evidence base. For instance, 2 MR papers^122,123^ and a meta-analysis of 16 RCT^123^ studies reveal no significant effect of vitamin D on blood pressure in the general population, with the exception of one MR study^124^ that indicated a blood pressure-lowering effect of higher vitamin D status. Similarly, a range of study types: one MR analysis^125^, one RCT^126^ and a meta-analysis of 27 prospective cohorts^127^ (only some of them RCTs) confirm a negative impact of smoking on HDL-C levels, bar one MR study showing no significant effect^128^. A series of RCT meta-analyses^129–131^ support an effect of coffee consumption (especially unfiltered) on unfavourable blood profile, although this is likely explained by diterpenes^132,133^ rather than caffeine, as the latter shows evidence of cardioprotective effects^134^. However, only the recent biggest MR study^133^ to date found a significant effect of coffee consumption on LDL-C and total-C levels, unlike two previous smaller MR analyses^135,136^, which found a non-significant directionally-consistent relationship.

## Discussion

Our study highlighted that sparsity of data in the electronic databases seriously hampers the ability to automatically parse and compare results of MR and RCT studies. Released for the first time in 2000, ClinicalTrials.Gov is the most comprehensive resource for modern RCT (only <1,000 studies, out of ∼167,000 analysed RCTs were started before 2000). Nonetheless, we found that only 11% of all completed RCTs submitted their results to ClinicalTrials.Gov, with median trial start date in 2012. Despite 2007 legislation requiring submission of RCT results to ClinicalTrials.gov within 1 year of completion (with exceptions)^137^, only 38% of eligible trials for 2008-2012 submitted their results at any time^138^ which rose to 64% for 2018-2019^139^. Furthermore, 60% of studies for failed agents are reported not to be published in peer reviewed journals^140^, and in the work presented here we found MeSH annotations were missing from the majority of complex, behavioural and dietary interventions. These factors significantly hamper efforts to systematically triangulate RCT evidence with other studies.

Next, semantic triples describing conclusions of MR and RCT studies automatically extracted from literature abstracts using rule-based methods also had low coverage, with only 25% of MR and 12% of RCT studies associated with >=1 triples. Consequently, we instead decided to focus on a detailed qualitative investigation of a series of case studies to identify the issues associated with triangulating MR and RCT studies

Combining RCT and MR results can offer complimentary evidence on the effectiveness of interventions. Powerful examples include congruence of positive effect of high BMI on hypertension across MR^100–103^ and RCT^92,93^ studies, high BMI on T2D risk in MR^82–86,104^ and RCTs^87–89^, and the null effect of vitamin D on various glycemic markers in diverse populations in MR^105^, RCTs^106,107^ and RCT systematic review^108^. We also found cases, where the majority of studies pointed to one direction of effect, with one MR or RCT identified as an outlier study. In these cases, having a wide array of MR and RCT studies (ideally meta-analysed) can be helpful in establishing the likely true causal direction of effect.

Our analysis of genetically predicted effects of perturbation of drug target protein expression on a number of conditions with trials submitted to ClinicalTrials.Gov revealed good concordance with established therapeutics for pQTLs. However, due to the limited number of proteins (n=1,002) and phenotypes (n=225, many non-diseases *per se*) in Zheng et al. (2020)^141^, the comparison is necessarily very preliminary. We identify only true positive cases, as false positives and true negatives are difficult to evaluate due to sparsity of drug clinical trial results in ClinicalTrials.Gov/literature^140^ and inclusion of non-disease phenotypes in MR analysis. Anecdotally, we found no MR evidence that decreased expression of PLA2G2A leads to reduced cardiovascular disease, which agrees with lack of efficacy of PLA2G2A inhibitor in clinical trials^142–144^.

The mixed reliability of eQTL instruments in predicting direction of effect on the outcome could be due to a number of factors such as: less than perfect correlation between mRNA and protein levels^145^, hidden pleiotropy in single instruments used in the MR analysis (directly observed for IL2RA)^146^, presence of negative feedback loop involved in the drug mechanism^147^, translation into protein isoforms with distinct biological effects^148^ and differential cell-type specific drug effect^149^.

The duration of intervention varies between RCTs and MR studies, with the former spanning no more than the duration of the trial, whilst the latter can represent durations as long as the entire lifetime (although many exposures, such as alcohol intake, will be over a shorter time period)^7^. Moreover, intervention in RCTs with long follow-up is not necessarily similarly intensive throughout its duration, or may cease altogether after some time^67,92^, i.e. duration of follow-up is longer than duration of intervention in order to allow accumulation of enough events and/or confirm durability of intervention effect. Examples include lifestyle interventions, such as exercise^67^ or weight loss programmes^87^ like the Look AHEAD trial, with median follow-up of 9.6 years, where group and individual counselling sessions took place weekly in the first 6 months and tapered off over time^66^. That is why our analysis focussed on comparing direction of effect, while ignoring magnitude of effect^150^. However, in certain cases when enough reference data is available, it is feasible to compare MR and RCT effects on the same exposure difference scale^151^.

Further impediments to direct comparison between MR and RCTs include differences in outcome definition (composite^58^ versus single conditions^63^). Access to rare subpopulations with existing conditions, such as cancer patients receiving specific therapy^152^ which are routinely exclusively enrolled into RCTs can be difficult in MR due to the size of GWAS biobanks relative to *N* required for good power.

There are also a number of interventions and outcomes with no single phenotype which could be instrumented with GWAS variants, making MR approaches difficult, although sometimes possible with innovative MR approaches^153^. This is especially true of lifestyle interventions – such as different forms of psychological therapy, complex diet regimens^154^ and fasting. Absent or limited heritability of a number of interventions and conditions, such as rehabilitation and traumatic injury makes MR approaches inaccessible.

The majority of MR studies track the onset rather than progression of disease due to availability of GWAS phenotypes^155^ which are often a (binary) single measurement, as opposed to multiple quantitative outcomes frequently measured in RCTs^156^. For that reason, triangulation of MR of onset with RCTs whose intervention is targeting progression of disease, may or may not result in agreement, as seen in our comparison of the effect of exercise on schizophrenia onset/progression (discordant) or depression (concordant) and vitamin D effect on atopic dermatitis onset/progression (discordant).

Many MR studies may be underpowered due to large sample required in indirect estimation^157^ as these studies are typically studies of convenience. This bias is less common in RCTs due to pre-registration of study design including power analysis^1^, uncommon in MR^158^. Null effect in MR studies may be therefore spurious and not predictive of RCTs for that reason, as seen in two smaller MR studies^135,136^ out of three^133^ investigating the effect of coffee intake on blood lipids, contrasting with strong clinical trial^129–131^ and biochemical evidence^132,133,159^.

Furthermore, the presented literature survey used a simple heuristic of reported statistically significant evidence (*p*-value < 0.05 after multiple testing correction) to compare conclusions across MR and RCT studies, which has well-known limitations^160,161^. Inclusion of the full-spectrum of scaled point-estimates along with their confidence intervals will reveal a more detailed picture in triangulation of MR and RCT evidence (Supplementary Box).

Our research highlights the challenges and benefits of triangulation of MR with RCT evidence. Future efforts, outside of the scope of this work, will focus on fully quantitative approaches towards triangulation, involving magnitude of effect size and not just its presence and direction^25^. Developers of such methods will need to be mindful of discrepancies in research hypothesis, duration and intensity of exposure, outcome measures, intervention aim, underlying population characteristics, violations of test assumptions as well as statistical power of the analysis. Furthermore, automated triangulation based on electronic databases requires intensive effort towards structured capture of both MR and RCT study results and associated meta-data, as well as annotation with shared ontologies, which is still challenging using current natural language processing methods, despite constant progress^36,162,163^.

## Methods

### ClinicalTrials.Gov *Data sources*

All ClinicalTrials.Gov study data is available for download as PostgreSQL database from The Database for Aggregate Analysis of ClinicalTrials.gov (AACT)^164^ released by The Clinical Trials Transformation Initiative^165^. We downloaded its static release from 1^st^ August 2021. Processing of the database files was carried out using custom Python and R scripts.

### ClinicalTrials.Gov *Data filtering*

We filtered the ClinicalTrials.Gov studies using a number of criteria to identify RCTs with submitted results allowing direct comparison with MR studies. These are depicted in Figure 2 and provided in detail in Supplementary Note.

### EpigraphDB *queries*

EpigraphDB^28^ was used to collect information about confirmed drug-target associations which were initially sourced from the Open Targets Platform^166^ and verified in DrugBank^167^. We also used EpigraphDB^28^ to source SemMedDB^35^ semantic triples associated with select MR and RCT publications identified by PubMed. SemMedDB triples in EpigraphDB are prefiltered for annotation of epidemiological studies as described previously^168^.

### PubMed *Data harvesting*

We searched PubMed for all RCT and MR studies published before 2021 on 1^st^ October 2021. For RCTs, we searched titles and abstracts for keywords: “randomized controlled trial” or “RCT” and we used PubMed’s in-built *Randomized Controlled Trial* label filter to obtain more specific hits, reducing the number of hits from 112,015 to 63,187. In order to retrieve potential MR studies, we used the keywords: “mendelian randomization” or “mendelian randomisation”. We also considered using MeSH labels “Randomized Controlled Trial” and “Mendelian Randomization Analysis” but both in case of RCT and MR studies they returned unrealistically low number of hits (7,999 and 1,926, accordingly), a consequence of manual indexing.

### Literature searches

We used Semantic Scholar and Google Scholar to survey MR and RCT literature indexed before 1^st^ August 2021. We queried the databases with the following search terms: “[exposure] [condition] Mendelian Randomization” and “[exposure] [condition] Randomized Controlled Trial”. The articles were initially screened by title and abstract. We included original research MR, RCT studies as well as meta-analyses. As exposures, we chose common lifestyle risk factors for chronic disease^169^ – body mass index (BMI), smoking, alcohol intake, blood pressure. Examples of dietary intervention (vitamin D/E, coffee) and behavioural intervention (exercise) were also selected. Outcomes included a number of cardiovascular disease and risk biomarkers, glycemic traits, neuropsychiatric disease, bone mineral density, atopic dermatitis and prostate cancer, amongst others (Supplementary Dataset 4). MR and RCT studies were compared across: population characteristics (sex, ethnicity, age, health status), comparator group, goal of intervention (prevention or treatment/slowing progression), direction of effect, length of follow-up, main test statistic in the study and its impact as judged by citation number.

## Supporting information

Supplementary Box

## Data Availability

All data produced in the present work are contained in the manuscript and supplementary files.

## Acknowledgments

We would like to acknowledge UK Medical Research Council [mc_uu_00011/4] which provided funding for our work carried out at the University of Bristol MRC Integrative Epidemiology Unit.

## Author contributions

TRG conceived and supervised the study as well as acquired funding. MKS curated the data and performed the main analyses. JZ inspired the study and supplied xQTL analyses. MKS, GDS, JZ and TRG wrote the manuscript.

## Data availability

Code used to carry out the analysis is available on GitHub: https://github.com/marynias/mr-rct. ClinicalTrials.Gov data was accessed via AACT: https://aact.ctti-clinicaltrials.org/snapshots and analysed data subset is available in Supplementary Datasets 1 & 2. pQTL and eQTL MR analysis results are available via EpigraphDB: https://epigraphdb.org/xqtl

## Supplementary Files

### Supplementary Box

#### Supplementary Note

##### Supplementary Figures

**Figure S1. Overview of RCT general features in the *Main dataset* by their frequency:** a) intervention model, b) primary purpose, c) trial status, d) trial phase, e) gender, f) number of arms.

**Figure S2. Distribution of: a) primary outcome number among RCT studies, b) secondary outcome number among RCT studies, c) P-value among study results**.

**Figure S3. Overview of SemMedDb semantic triples derived from MR (blue, left-hand panels) and RCT (orange, right-hand panels) studies:** a) & b) top 15 subjects, c) & d) top 15 predicates, d) & e) top 15 objects.

##### Supplementary Tables

**Table S1. Comparison of the most common intervention types in the *Main* and *Literature* datasets**.

**Table S2. Drug target-disease matches supported by evidence from MR (blood eQTL instruments**, https://epigraphdb.org/xqtl**) and RCT studies (*Main dataset* from ClinicalTrials.Gov)**.

**Table S3. Matching subject-object pairs between MR and RCT studies in semantic triples identified by SemMedDb**.

**Table S4. Matching subjects between MR and RCT studies in semantic triples identified by SemMedDb**.

**Table S5. Matching objects between MR and RCT studies in semantic triples identified by SemMedDb**.

**Table S6. Sample comparison of MR and RCT studies with matching exposures and outcomes across a range of criteria**.

##### Supplementary Datasets

**Supplementary Dataset 1**. Select ClinicalTrials.Gov data fields for the RCT studies identified as the *main dataset*.

**Supplementary Dataset 2**. Select ClinicalTrials.Gov data fields for the RCT studies identified as the *literature dataset*.

**Supplementary Dataset 3**. Comparison of frequency of RCT general features between the *Main dataset* and background of all RCTs available in ClinicalTrials.Gov.

**Supplementary Dataset 4**. Case series of MR and RCT studies with matching exposures (interventions) and outcomes (conditions).

