## Supplementary figures and images for "Systematic comparison of Mendelian randomization studies and randomized controlled trials using electronic databases"

### Supplementary Figure S1

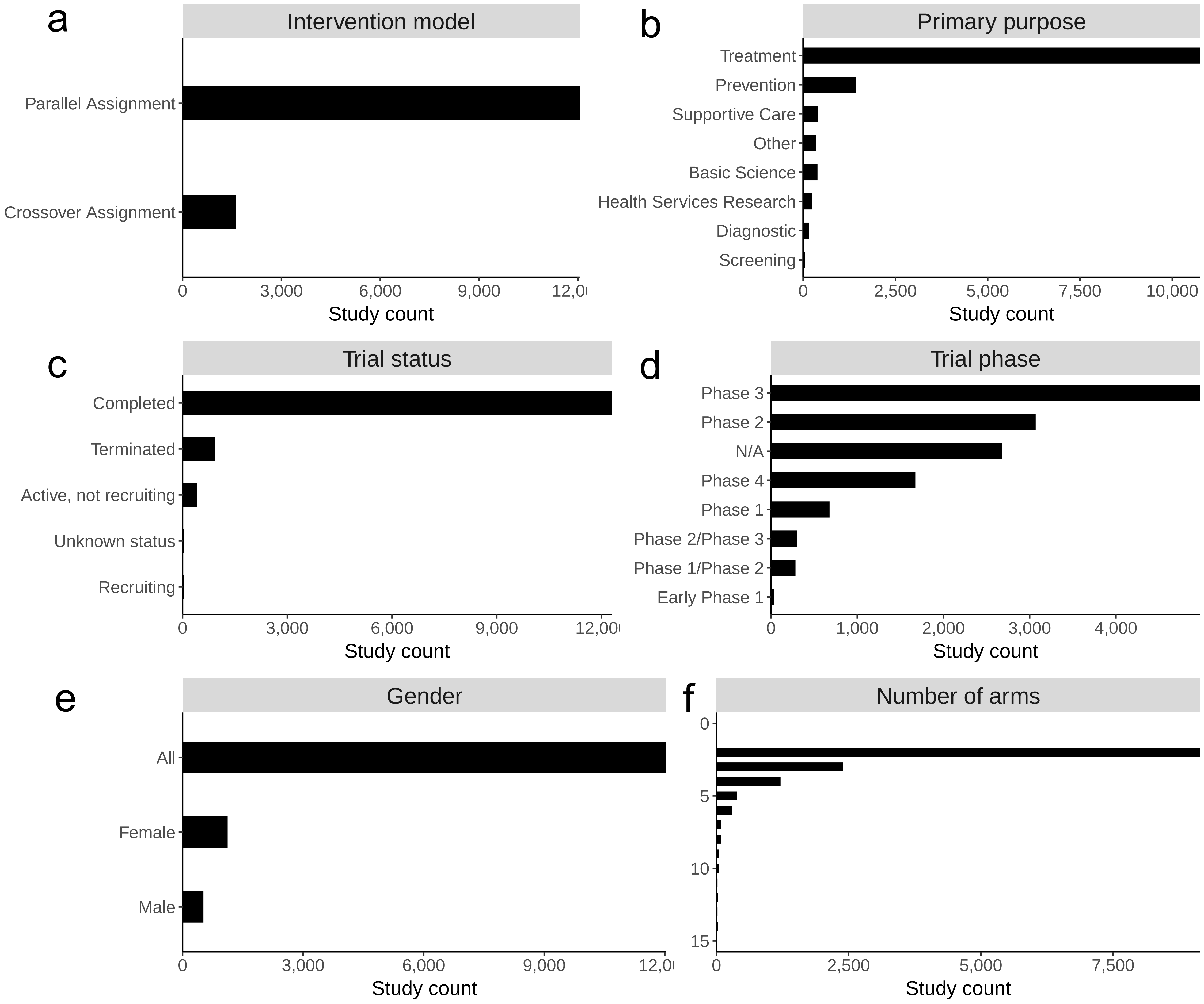

### Supplementary Figure S2

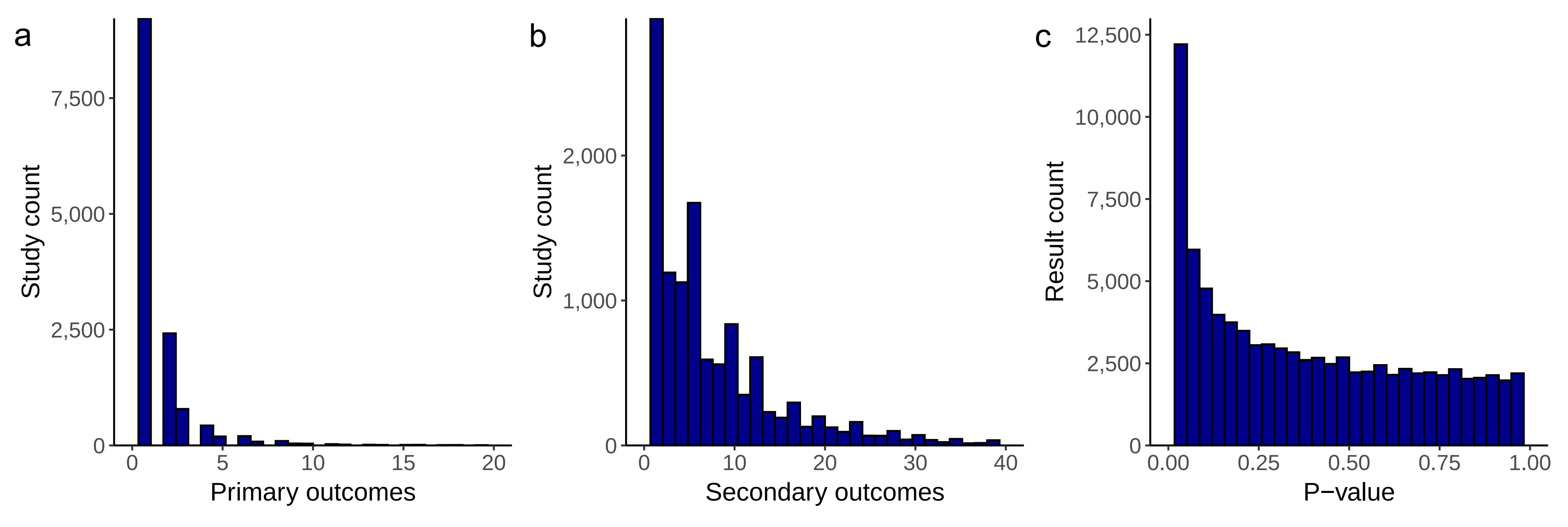

### Supplementary Figure S3

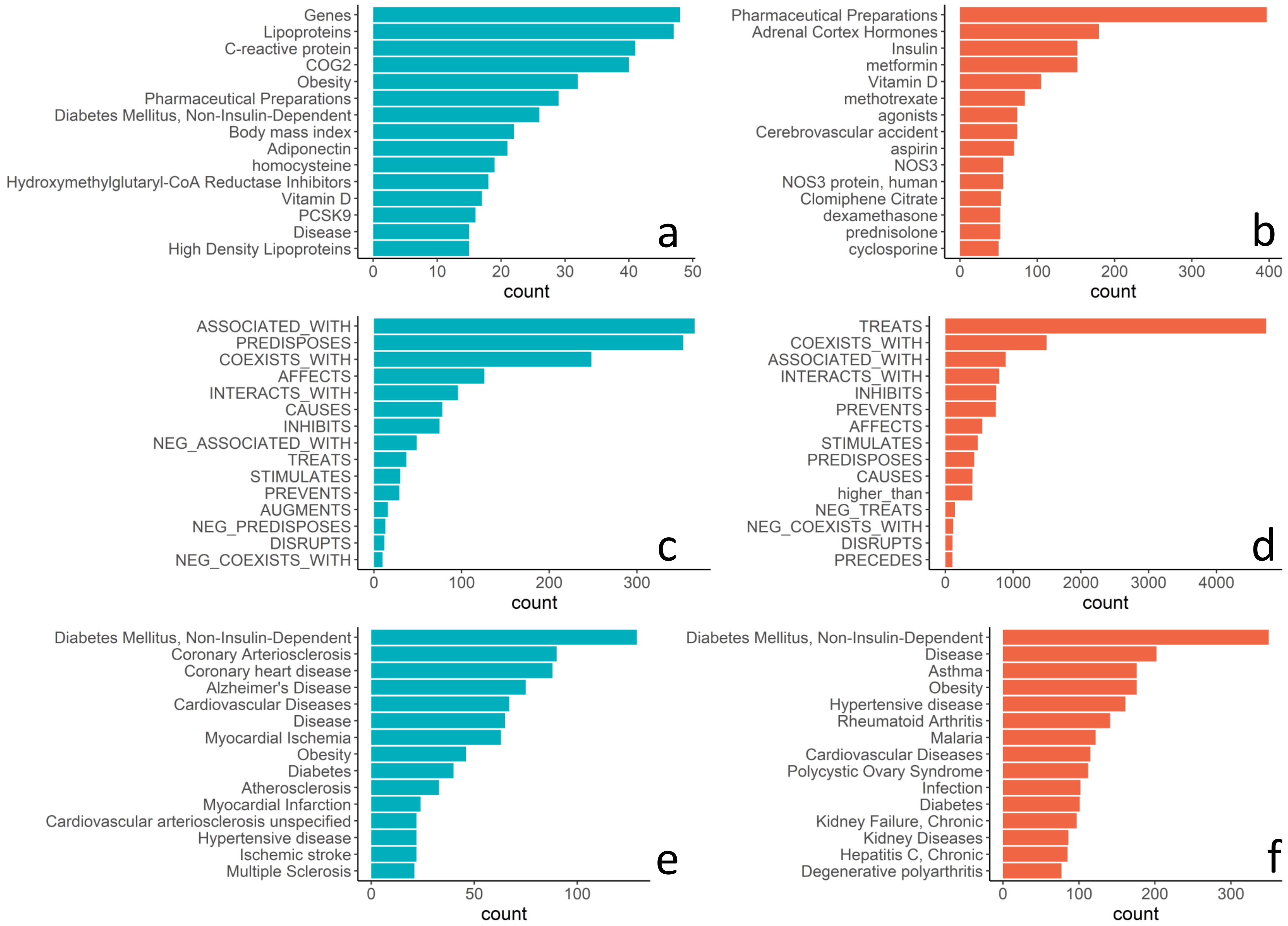
