## Supplementary Note for "Systematic comparison of Mendelian randomization studies and randomized controlled trials using electronic databases"

ClinicalTrials.Gov *Data filtering*

We only included studies with most common designs suitable for comparison with published MR studies: *Parallel Assignment* and *Crossover Assignment* (eliminating: *Single Group Assignment, Sequential Assignment, Factorial Assignment*) intervention model in the Designs table with *Randomized* allocation to allow for selection of RCT. We further used the study type field (=*Interventional*) with a minimum number of arms = *2* in the Studies table as an additional filtering criterion to arrive at a set of RCT studies. The next stage of filtering concerned background information and study results. We first filtered on the presence of a study description in the Brief Summaries table. The key criterion was then presence of results in the Outcome Analyses table, where we selected the variables: *param_type, param_value, p_value* and *method.* Next, we needed all conditions to have at least 1 Medical Subject Headings (MeSH) term assigned in all_conditions view to facilitate automatic comparison with external data sources. We dropped that requirement for interventions, as especially behavioural interventions could not be assigned a MeSH term (see Results). Finally, a range of basic reference and eligibility criteria were required: *brief_title, study_type, overall_status, phase, number_of_arms, enrolment* in the Studies table, *gender, criteria* in the Eligibilities table and *outcome title* and *type* in the Outcomes table.

Additionally, we extracted a subset of studies which did not supply any results in the Outcome Analyses table and therefore did not contribute to the *Main* dataset above but were RCT studies with published literature records in the Study References table (*reference type* = result).

*Overview of top SemMed triples for MR and RCT studies*

An overview of the top subjects in MR (Figure S3a) and RCTs (Figure S3b) revealed a high number of terms related to adiposity (*obesity, body mass index, adiponectin*), lipid biology (*lipoproteins, hydroxymethylglutaryl-CoA reductase inhibitors, PCKSK9, high density lipoproteins*) as well as *type 2 diabetes* and *vitamin D*. Terms related to type 2 diabetes (*insulin, metformin*) and *vitamin D* were also found in RCT triples. As expected by the preponderance of drug interventions in ClinicalTrials.Gov, there was a noticeable bias towards pharmaceutical preparations amongst the top 10 subjects in RCT triples with terms such as *methotrexate*, *aspirin*, *clomiphene citrate*, *dexamethasone*, *prednisolone* and *cyclosporine.* SemMedDb identified *associated with* as a top predicate in MR studies (Figure S3c), followed by *predisposes*, *coexists with* and *affects* which underlines the skew of current MR studies towards identifying risk factors for disease. On the other hand, RCT studies lean towards identifying treatments (Figure S3d) which is reflected in the top 1 predicate *treats* (*n*= 4,732, second-best *coexists with* n= 1,495). Among the top objects in MR studies (Figure S3e), we found cardiovascular diseases (*coronary arteriosclerosis*, *coronary heart disease*, *myocardial ischemia/infarction*, *hypertensive disease*, *ischemic stroke*). High frequency of a smaller number of cardiovascular disease terms (Figure S3f) was also found among RCT objects (*hypertensive disease, cardiovascular disease*). Both MR and RCT showed type 2 diabetes and obesity as commonly studied conditions. RCT objects included also two infectious diseases: malaria and hepatitis C.
