## Supplementary Box for "Systematic comparison of Mendelian randomization studies and randomized controlled trials using electronic databases"

It is anticipated that the longer duration of exposure differences seen in MR studies will generate larger effect sizes than will relatively short-term modification of the exposure in RCTs. For example, the genetic variants related to non-HDL cholesterol (henceforth “cholesterol”, the target of cholesterol lowering drugs such as the statins) have been shown to relate to relatively stable differences from early childhood to late adulthood, thus generating a lifetime of differential exposure to circulating cholesterol. The RCTs of cholesterol lowering drugs designed to show effects on coronary heart disease (CHD) events last ~5 years. Atherosclerosis is a disease process that develops from childhood onwards, and the CHD it generates would not be expected to be abolished by a few years of cholesterol lowering in middle age or older (the usual age included in the RCTs). As there are several cholesterol lowering drugs which target different genes it is possible to compare MR studies using genetic variants in those genes that are robustly related to cholesterol with RCTs of drugs that target those genes. Figure SB1 below combines data from such MR analyses with the RCTs.

As anticipated the RCTs produce about 40% of the risk reduction seen with a lifetime difference in exposure levels^1^. This scaling of the effects predicted from MR studies and seen in the matching trials can be applied to MR/RCT comparisons for other exposures, as the time course of effects being produced may be quicker than seen in the case of cholesterol, or take longer, or indeed there may be no effect in the RCTs if the effect of the exposure acts during a critical period in earlier life and sets in train a disease process that is not reversible by later modification of the exposure^2^.

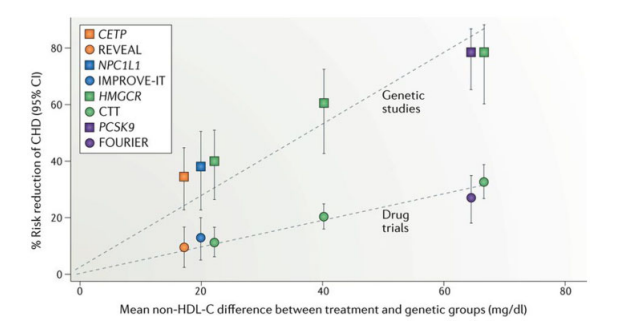
**Figure SB1.** Drug treatment (circles) and genetic proxy (squares) effects on reducing cholesterol levels and the corresponding reduction in risk of from matching drug RCTs and MR analyses. The colours indicate the gene from which variants are taken in the MR studies and the target of the drug used in the named RCTs. *Abbreviations: CETP, cholesteryl ester transfer protein; HMGCR, 3-hydroxy-3-methylglutaryl-CoA reductase; NPC1L1, Niemann-Pick C1-like protein 1; PCSK9, proprotein convertase subtilisin/kexin type 9.* Figure reproduced with permission from^3^.
